# Low testosterone levels in H7N9 infected men correlate with high cytokine responses and lethal outcome: a retrospective cohort study

**DOI:** 10.1101/2020.05.07.20093914

**Authors:** Yongkun Chen, Tian Bai, Sebastian Beck, Stephanie Stanelle-Bertram, Tao Chen, Jie Dong, Jing Yang, Lijie Wang, Dayan Wang, Yuelong Shu, Gülsah Gabriel

**Author notes:** equally contributed first authors (Yongkun Chen, Tian Bai, Sebastian Beck). equally contributed senior authors (Yuelong Shu, Gülsah Gabriel). Correspondence to: Gülsah Gabriel, PhD or Yuelong Shu, PhD.

## Abstract

**Background:** Human infections with avian influenza A (H7N9) virus emerged in East China in March 2013. In contrast to seasonal influenza A viruses, H7N9 infections present a strong sex bias. Over the five epidemic waves in China, ~70% of all H7N9 cases were observed in men. We and others have independently shown that influenza A virus infections may significantly lower testosterone levels in male mice. In this study, we sought to analyze the role of testosterone in disease outcome in H7N9 infected men.

**Methods:** We systematically analyzed a total of *n*=330 human samples obtained from male and female subjects of two age groups (18-49 years and ≥50 years) with laboratory-confirmed H7N9 influenza, seasonal influenza (H1N1, H3N2) or negative control cohorts including H7N9 virus-negative close contacts and influenza virus-negative poultry workers in dependency of sex. The level of testosterone, estradiol as well as cytokines and chemokines were measured and compared in all study participants. We used regression analysis to evaluate the association of sex hormones and cytokines in dependency of sex and disease outcome.

**Results:** We analyzed sex hormones and innate immune responses in H7N9 cases (*n=*98), their H7N9 virus-negative close contacts (*n=*71), influenza virus-negative poultry workers (*n=*108) and mild seasonal (H1N1, H3N2) influenza cases (*n=*53). Samples were collected between 2014 and 2017. All control groups showed a similar median age within H7N9 cases except for the seasonal influenza group with a younger median age. In H7N9 infected men, testosterone levels were strongly reduced compared to male H7N9 virus-negative close contacts or males with seasonal influenza. Low testosterone levels in H7N9 infected men correlated with high inflammatory cytokine levels, e.g. IL-6, and lethal outcome in 18-49 year olds. No significant differences were detected in estradiol levels in H7N9 infected men. In H7N9 infected women (≥ 50 years), estradiol levels were elevated compared to H7N9 virus-negative close contacts without significantly affecting disease outcome.

**Conclusions:** Here, we show that low testosterone levels pose a poor prognostic marker in H7N9 infected men. Thus, treatment of H7N9 infected patients should consider sex-specific mitigation strategies.

**Key point:** Avian H7N9 infection presents a higher incidence in infected men compared to women. Low testosterone levels in H7N9 infected men correlate with high-level cytokine expression posing a high risk factor for lethal outcome.

## Introduction

In early 2013, human infections with avian influenza A (H7N9) virus were first reported in China[1]. Since then, the H7N9 avian influenza A virus caused five epidemic waves from 2013 to 2017. Exposure at live poultry markets was considered a major risk factor for human infections with H7N9 virus. A primary epidemiological study revealed that 71% of all H7N9 cases were males in the first wave [2]. The high degree of sex bias in men consistently accounted with similar proportions (68%-71%) during the entire five H7N9 epidemic waves[3]. Currently, very little knowledge exists on the complex interplay of biological sex and gender specific behavior as contributors to influenza disease outcome. The World Health Organization (WHO) Western Pacific team proposed gender-associated practices and norms, such as more frequent exposure of men to birds, may pose one of the reasons for sex-specific H7N9 incidence[4]. However, a follow-up study suggested increased risk in older men is not due to higher exposure time to live poultry market[5].

Sex hormones, particularly testosterone and estradiol are important biological factors that affect sex differences in immune responses [6–9]. We have recently shown that H1N1 infection itself reduces testosterone levels in male but not female mice. Conversely, treatment of female mice with testosterone protected the majority of H1N1 infected females from lethal outcome[10]. In line, aged male mice with low testosterone levels were reported to undergo elevated pulmonary inflammation and severe disease upon H1N1 influenza virus infection compared to young male mice[11]. Conversely, other murine models showed that low concentrations of estradiol in female mice was associated with enhanced pro-inflammatory responses and H1N1 influenza pathogenesis [12]. In contrast, high estradiol levels protected female mice from lethal influenza infection by dampening inflammatory cytokine responses [12]. Thus, there is increasing evidence from small animal models that sex hormones play an important role in influenza disease outcome.

However, very little knowledge exists on the role of sex hormones in infectious diseases in humans and their association with disease outcome. Thus, in this study, we retrospectively analyzed the impact of sex hormones on influenza disease outcome in males and females.

## Materials and Methods

### Ethics statement

Sampling from laboratory-confirmed H7N9 avian influenza cases, seasonal influenza cases, close contacts of H7N9 cases and poultry workers was reviewed and approved by the Ethics Committee of the National Institute for Viral Disease Control and Prevention, China CDC.

### Study participants

Stored serum samples from acute phase of laboratory-confirmed H7N9 hospitalized patients were collected from 2014 to 2017 through the influenza surveillance network in the local Center for Disease Control and Prevention (CDC), China. H7N9 patients (*n=*98) aged above 18 years with complete epidemiological information (age, sex, illness onset date, sample collection date, outcome and antiviral treatment) and required amount of serum were enrolled in this study. Once a patient was confirmed with H7N9 influenza virus, blood sample and throat swabs of the close contacts and epidemiological linked poultry workers were collected through the influenza surveillance network of the local CDC, China. Swabs were first screened by real-time RT-PCR to identify influenza A and B viruses in the local CDC or the Chinese National Influenza Center. Positive samples for influenza A virus were further tested for influenza A (H7N9) virus. No influenza A (H7N9) virus was detected from close contacts and poultry workers. A total of 71 close contacts and 108 poultry workers were included in this study as control groups by matching age or sex of H7N9 cases. Archived plasma samples from laboratory-confirmed outpatients infected with influenza A (H1N1) pandemic 2009 or influenza A (H3N2) viruses were collected from 2015-2016 influenza season in China. Plasma samples and throat swabs were collected simultaneously when the patient went to outpatient care. Blood samples were archived when seasonal influenza virus was detected by real-time RT-PCR. Seasonal influenza cases (2009 pandemic H1N1: *n=*22, H3N2: *n=*31) were selected as another control group in this study. All samples from study participants were stored at -80 °C until further analysis.

### Hormone quantification

The total concentration of estradiol (17β-estradiol) and testosterone was measured in the clinical laboratory of the Sino-Japan Friendship Hospital (Beijing, China) from collected serum or plasma samples of all study participants using the Beckman Coulter DxI 800 Access Immunoassay System. The majority of circulating estradiol and testosterone are bound to the sex hormone binding globulin (SHBG), but it also exists loosely bound to albumin and in the free status. We used the Access Estradiol Assay (REF 33540, BECKMAN COULTER) and Access Testosterone Assay (REF 33560, BECKMAN COULTER) which use paramagnetic particles in a chemiluminescent immunoassay for the quantitative determination of total estradiol and testosterone level (bound and unbound).

### Cytokine and chemokine measurement

Serum or plasma samples of H7N9 and seasonal influenza cases were analyzed for a panel of 29 cytokines and chemokines (EGF, Eotaxin, G-CSF, GM-CSF, IFN-α2, IFN-γ, IL-10, IL12-P40, IL12-P70, IL-13, IL-15, IL-17A, IL-1RA, IL-1α, IL-1β, IL-2, IL-3, IL-4, IL-5, IL-6, IL-7, IL-8, IP10, MCP-1, MIP-1α, MIP-1β, TNF-α, TNF-β, VEGF) by Luminex bead based multiplex assay using human cytokine/chemokine magnetic bead panel 96 well plate assay (HCYT0MAG-60K, Millipore, USA) according to manufacturer’s instruction. A smaller panel including 21 selected cytokines and chemokines, where significant differences were observed earlier (GM-CSF, IFN-α2, IFN-γ, IL-10, IL12-P40, IL12-P70, IL-13, IL-17A, IL-1RA, IL-1α, IL-1β, IL-5, IL-6, IL-7, IL-8, IP10, MCP-1, MIP-1α, MIP-1 β, TNF-α, TNF-β) were analyzed for close contacts of H7N9 cases and poultry workers using the same measurements. For undetectable cytokines and chemokines (below the lowest detection limit) in all groups, we assigned a value as 0·1 for analysis.

### Statistical analysis

Statistical significance was determined with GraphPad Prism 5 v.5·03 (GraphPad Software, Inc.). Difference of sex hormones, cytokines/chemokines among multiple groups was analyzed using One-Way ANOVA (Kruskal-Wallis test) and selected Dunn’s as a post-test to compare each group. Since the results of cytokine/chemokines exhibited deviated scale among all groups, all values in the statistical analysis and figures were used after Ln-transformation. The difference of sex hormones and cytokines/chemokines between survival and dead H7N9 cases in each age group was analyzed using the unpaired, two-tailed Student’s *t*-test (non-parametric). Association between sex hormones and cytokine/chemokines were determined using linear regression and correlation analysis (Pearson). Since the results of cytokine/chemokines exhibited deviated scale than testosterone, all values in the statistical analysis and figures were used after Ln-transformation in comparison of cytokines/chemokines in all groups and linear regression with testosterone. Statistical significance was defined as *p* < 0·05 (* *p* < 0·05, ** *p* < 0·01, *** *p* < 0.001).

## Results

### Participant characteristics

A total of *n=*44 avian H7N9 influenza positive cases of reproductive age (18-49 years) were enrolled with a median age of 42 years (IQR 34-46 years) (Table 1). A total of *n=*54 avian H7N9 influenza positive cases were included in those ≥50 years old with the median age of 61 years (IQR 56-67 years) (Table 1). The male H7N9 cases accounted for 75% in the younger and 70% in the older age groups, which is consistent with previous epidemiological studies based on larger laboratory-confirmed H7N9 cohorts. Blood samples of H7N9 patients were collected within acute phases of 6·5 days (IQR 5-8·3 days) and 7·5 days (IQR 6-9 days) after illness onset. H7N9 cases confirmed in 2017 accounted for over 77% of all cases enrolled since 2014, in this study, presenting most of the participants from South China (data not shown). For all control groups, the median age is similar in close contacts and poultry workers compared to H7N9 cases in both younger and older age groups (Table 1). However, the age of seasonal influenza cases in younger age groups was lower compared to H7N9 cases. Consistent with H7N9 cases, the proportion of males in all control groups is higher than females, except for the seasonal influenza cases with a similar proportion in both sexes.

**Table 1:** Age and sex characteristics of study participants.

| Age group | Study participants | Age, years |  | Sex (%) |  |
| --- | --- | --- | --- | --- | --- |
|  |  | Median | IQR | Male | Female |
| **18-49 years** | Seasonal influenza cases(*n*=28) | 26.5 | 23.3-32.8 | 16 (57%) | 12 (43%) |
|  | Poultry workers (*n*=43) | 41 | 36-46.5 | 32 (74%) | 11 (26%) |
|  | H7N9 close contacts (*n*=40) | 35 | 27-41.3 | 29 (73%) | 11 (27%) |
|  | H7N9 cases (*n*=44) | 42 | 34-46 | 33 (75%) | 11 (25%) |
| **≥50 years** | Seasonal influenza cases (*n*=25) | 61 | 56-67 | 12 (48%) | 13 (52%) |
|  | Poultry workers (*n*=65) | 55 | 53-62 | 45 (69%) | 20 (31%) |
|  | H7N9 close contacts (*n*=31) | 62 | 54-70.5 | 16 (52%) | 15 (48%) |
|  | H7N9 cases (*n*=54) | 61 | 56-67 | 38 (70%) | 16 (30%) |

Data are median (Interquartile Range, IQR) or *n*(%).

### Low testosterone levels in H7N9 infected men are associated with lethal outcome

In order to assess the role of sex hormones in the observed sex bias of H7N9 cases, we first measured testosterone concentrations in all cohorts. Testosterone levels were strongly reduced in H7N9 infected men of both age groups assessed compared to virus negative H7N9 close contacts, poultry workers and those with seasonal influenza (Figure 1A). Low testosterone levels strongly correlated with lethal outcome in H7N9 infected men in the age group of 18-49 years olds (Figure 1B). No significant differences in testosterone levels were observed in H7N9 infected women compared to their close contacts and poultry workers. However, testosterone levels were significantly increased in those infected with seasonal influenza compared to all other female groups (Figure 1C). However, no significant impact of testosterone levels was observed regarding H7N9 disease outcome in women (Figure 1D). These data show that low testosterone levels in H7N9 infected men of 18-49 years of age correlate with an enhanced risk for lethal outcome.

**Figure 1:**
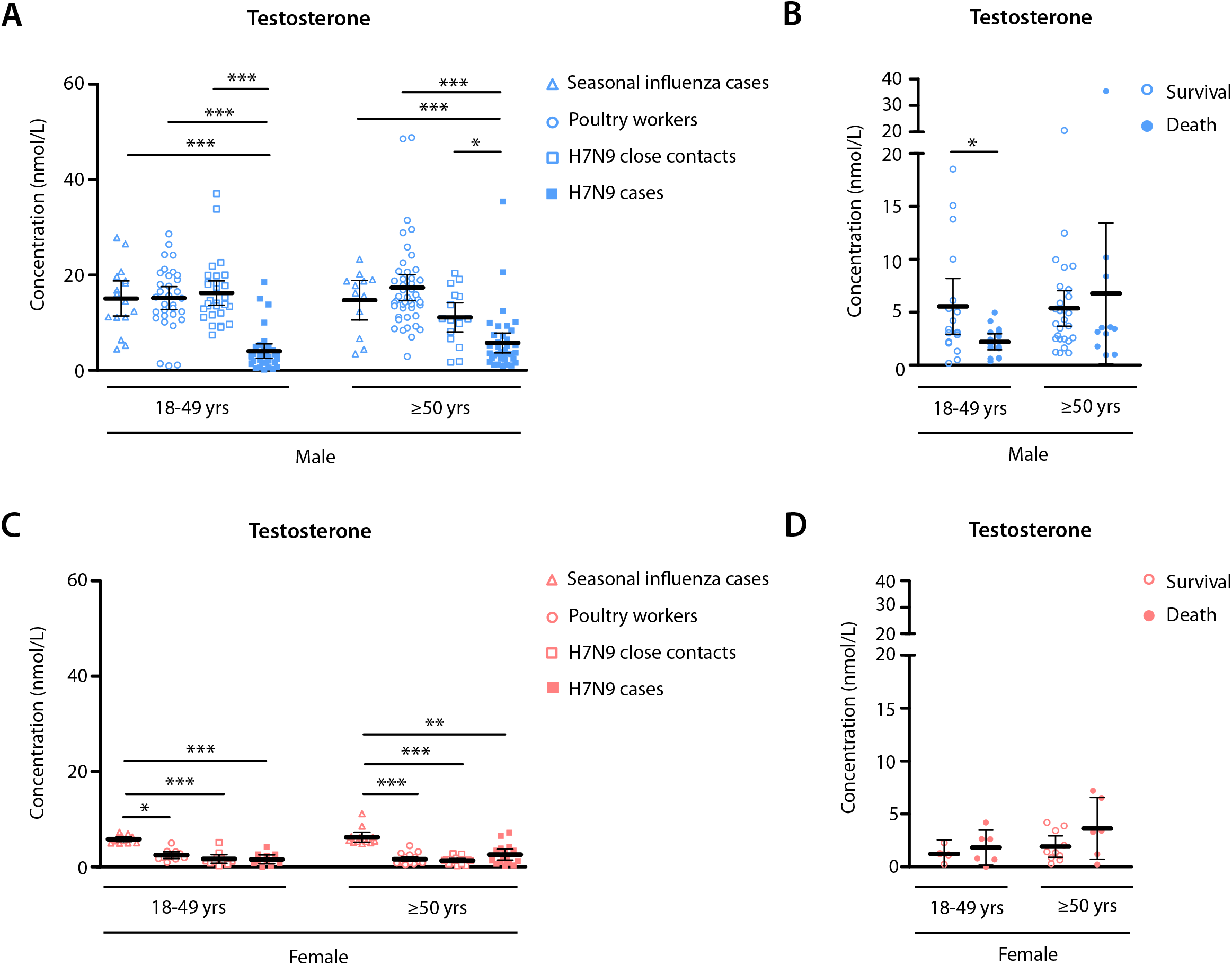
Testosterone expression levels in H7N9 IAV infected patients compared to control groups. (A, C) Total testosterone expression levels were measured in H7N9 IAV infected male (A) and female (C) patients (H7N9 cases, filled squares) and compared to control groups: i) seasonal influenza cases (triangles), ii) poultry workers (circles), and iii) H7N9 close contacts (open squares). (B, D) Total testosterone expression levels were measured in H7N9 IAV infected male (B) and female (D) patients (H7N9 cases) and shown in dependency of disease outcome (survival, open circles; death, filled circles). (A-D) Samples from both sexes were divided into two groups based on age: 18-49 and ≥ 50 years (yrs) old. Data represent the mean ± 95% confidence interval (CI). Numbers of samples are summarized in Table 1. Significant differences among all study groups were calculated using either one-way ANOVA (Kruskal-Wallis test, Dunn’s post-hoc-test) (A, C) or the unpaired, two-tailed Student’s *t*-test (B, D). Statistical significant differences were further classified into three groups: * p < 0·05, ** p <0·01, *** p < 0·001.

### High estradiol levels in H7N9 infected men is not associated with disease outcome

Conversely, we measured estradiol levels as a predominant female hormone in all cohorts. Seasonal influenza infected men of 18-49 years of age presented slightly but significantly increased estradiol levels compared to poultry workers, H7N9 cases and their H7N9 virus-negative close contacts (Figure 2A). No differences were detected among all groups of ≥50 years of age (Figure 2A). However, estradiol levels were not associated with disease outcome in H7N9 infected men (Figure 2B). In H7N9 infected women of ≥50 years of age, estradiol levels were significantly elevated compared H7N9 virus-negative close contacts (Figure 2C). Increased estradiol levels, however, were not significantly associated with disease outcome in females but a tendency towards an increased risk for lethal outcome may be speculated among both age groups (Figure 2D). These findings suggest that estradiol does not significantly affect disease outcome in H7N9 infected men.

**Figure 2:**
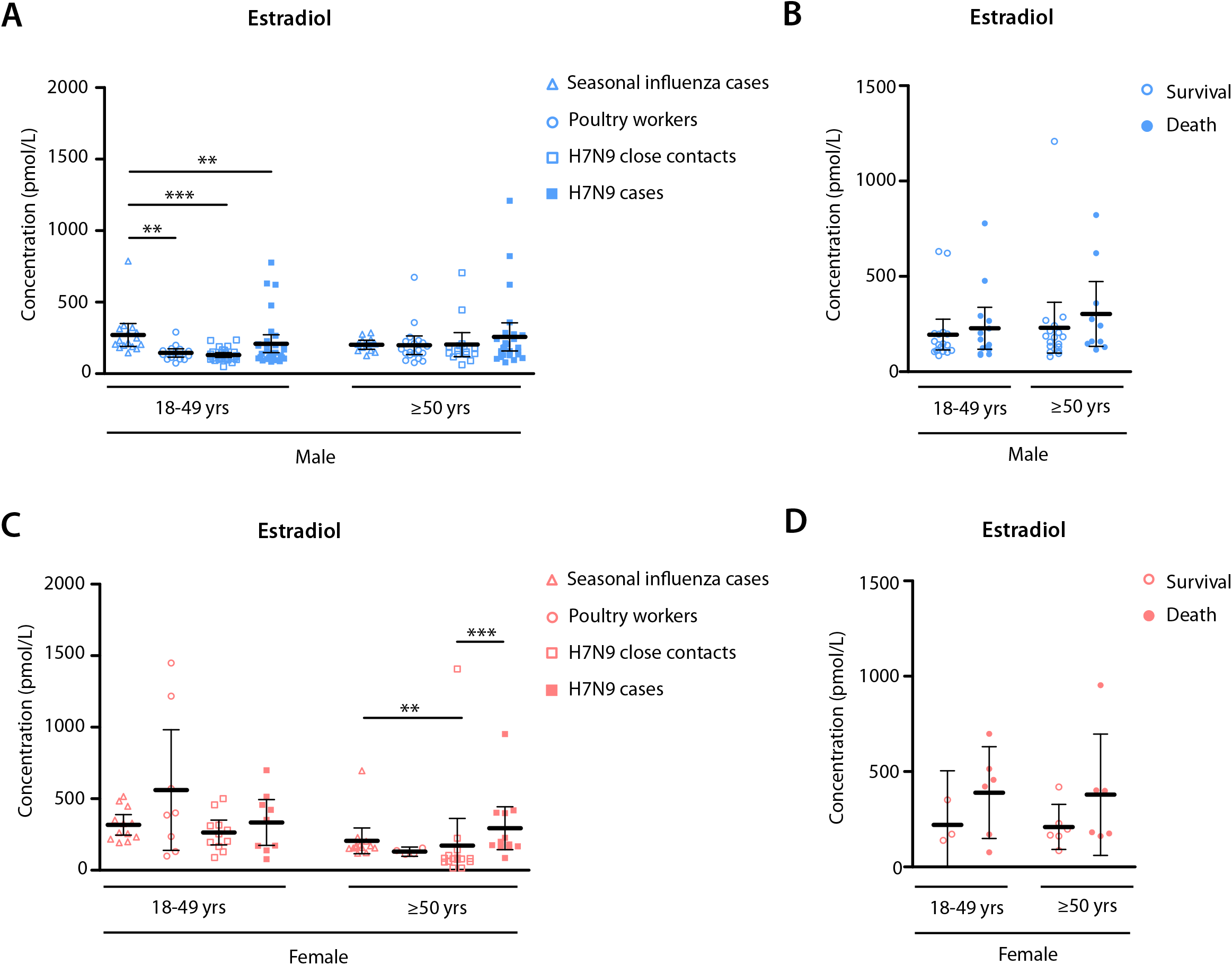
Estradiol expression levels in H7N9 IAV infected patients compared to control groups. (A, C) Total estradiol expression levels were measured in H7N9 IAV infected male (A) and female (C) patients (H7N9 cases, filled squares) and compared to control groups: i) seasonal influenza cases (triangles), ii) poultry workers (circles), and iii) H7N9 close contacts (open squares). (B, D) Total estradiol expression levels were measured in H7N9 IAV infected male (B) and female (D) patients (H7N9 cases) and shown in dependency of disease outcome (survival, open circles; death, filled circles). (A-D) Samples from both sexes were divided into two groups based on age: 18-49 and ≥50 years (yrs) old. Data represent the mean ± 95% confidence interval (CI). Numbers of samples are summarized in Table 1. Significant differences among all study groups were calculated using either one-way ANOVA (Kruskal-Wallis test, Dunn’s post-hoc-test) (A, C) or the unpaired, two-tailed Student’s *t*-test (B, D). Statistical significant differences were further classified into three groups: * p < 0·05, ** p < 0·01, *** p < 0·001.

### H7N9 infection induces a cytokine storm in men and women

Next, we assessed the cytokine and chemokine responses in H7N9 infected patients compared to their close contacts, poultry workers or those infected with seasonal influenza. In general, H7N9 infection resulted in highly increased cytokine and chemokine levels compared to their H7N9-negative close contacts or poultry workers (Figure S1 and S2). Seasonal influenza virus infected patients also presented elevated cytokine and chemokine levels but in general less than those infected with H7N9 (Figure S1 and S2). Thus, H7N9 induced cytokine storm is detected in men as well as women.

### Storm cytokine levels in H7N9 infected men are associated with lethal outcome

We then analyzed whether certain cytokines might be associated with differential disease outcome. In H7N9 infected men, elevated levels of G-CSF, GM-CSF, IL-8, IL-10, IL-15, MCP-1 and TNF-α were associated with death (Figure 3). In H7N9 infected women, elevated levels Eotaxin correlated with survival (Figure 3P), whereas elevated MIP-1ß (Figure 3R) and EGF (Figure 3T) levels were associated with death. However, direct comparison of cytokine and chemokine responses in H7N9 infected men and women revealed important differences (Figure S3). Lethal outcome in H7N9 infected men correlated with elevated Eotaxin and MIP-1ß levels in the 18-49 year olds and with elevated IL-7 in the ≥50 year olds unlike in age-matched women with H7N9 influenza (Figure S3A-C). Conversely, survival in H7N9 infected men correlated with elevated IL-7 and MIP-1ß levels (Figure S3B and S3C). These findings suggest that differential cytokines are associated with disease outcome in males and females.

**Figure 3:**
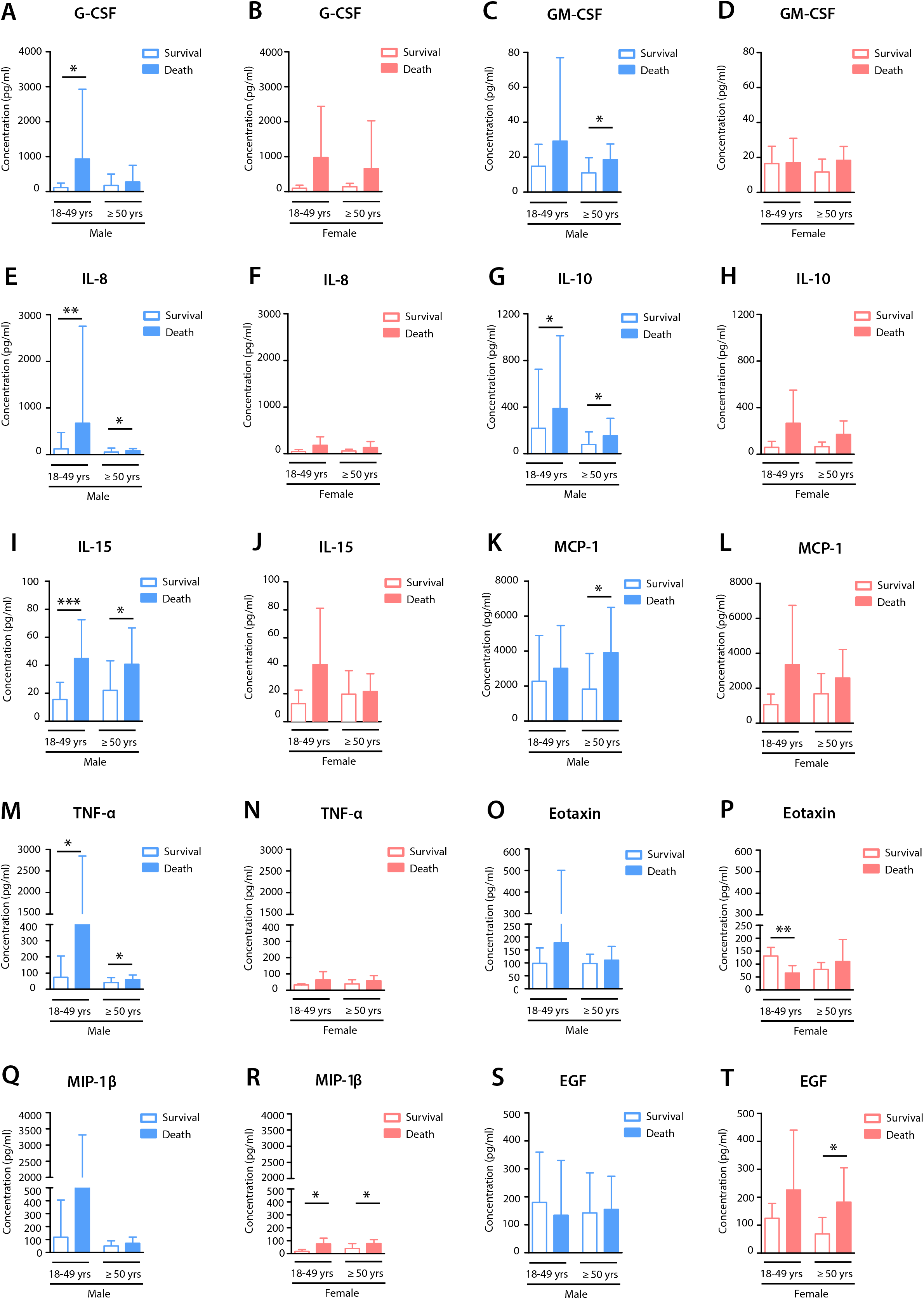
Cytokine and chemokine expression levels in H7N9 IAV infected patients in dependency of disease outcome. Shown are the significant expression levels of cytokines and chemokines in the sera of H7N9 IAV infected male and female patients who either survived or succumbed to the infection (survival and death, respectively). Samples were divided into two groups based on age: 18-49 and ≥50 years (yrs) old. The measurement was carried out using a multiplex immunoassay. Analytes included were: G-CSF (granulocyte-colony stimulating factor), GM-CSF (granulocyte-macrophage colony-stimulating factor), IL-8 (interleukin 8), IL-10 (interleukin 10), IL-15 (interleukin 15), MCP-1/CCL2 (monocyte chemoattractant protein 1), TNF-α (tumor necrosis factor alpha), eotaxin, MIP-1β/CCL4 (macrophage inflammatory protein 1 beta), EGF (epidermal growth factor). Values are shown as means ± SD. Statistical significant differences between groups were determined using the unpaired, two-tailed Student’s *t*-test and divided into: * *p* < 0·05, ** *p* < 0·01, *** *p* < 0·001.

### Low testosterone levels correlate with cytokine storm in H7N9 infected men

We could show that (1) high cytokine levels and (2) low testosterone levels correlate with lethal outcome in H7N9 infected men. In order to assess potential mechanistic links between individual cytokines and testosterone, we compared their individual levels in the same patient with disease outcome. In H7N9 infected men (18-49 years old), among all assessed cytokines, particularly IL-6, IL-8, IL-10, IL-1RA and GM-CSF levels were negatively associated with testosterone concentrations in H7N9 survivors with the exception of TNF-α. Thus, a significant number of H7N9 surviving men had high testosterone and low cytokine levels (Figure 4A-E). In contrast, all men who succumbed to H7N9 influenza presented low testosterone and high cytokine levels. In H7N9 infected men ≥50 years, a similar correlation was observed for the same cytokines as described for the 18-49 years olds (Figure 4 G-L). However, death was not significantly associated with low testosterone and high cytokine levels. In H7N9 infected women, positive linear association between cytokine and testosterone levels was observed unlike the observations in men (Figure S4). These findings, suggest that in men, expression levels of cytokines, such as IL-6, increases with reducing testosterone levels suggestive of potentially interlinked mechanisms.

**Figure 4:**
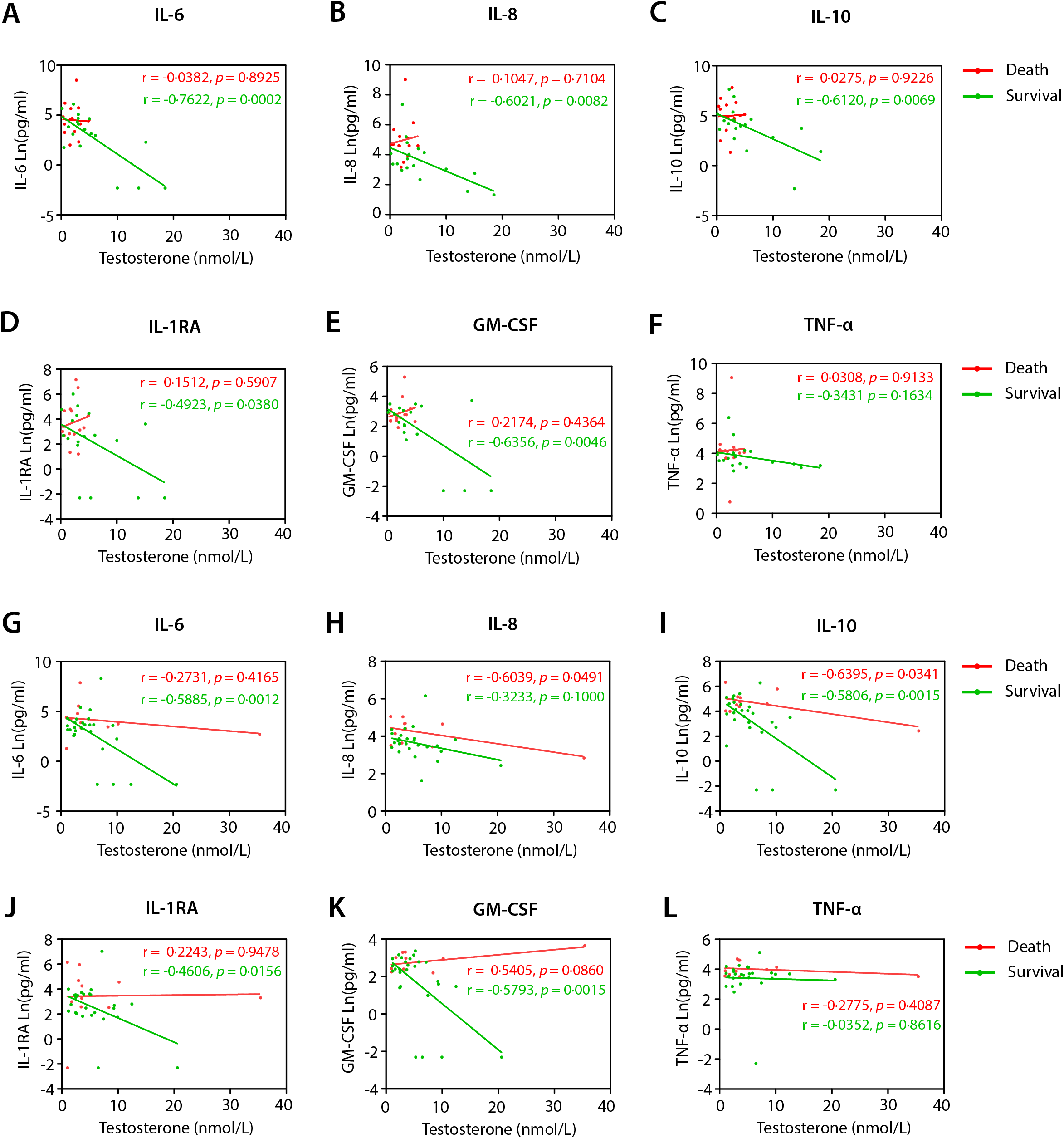
Correlative analysis of testosterone-modulated inflammatory immune responses in H7N9 IAV infected male patients. Testosterone expression levels measured in the sera from H7N9 IAV infected male patients (A-F, age group 18-49 yrs old; G-L, age group ≥ 50 yrs old) were plotted over the expression levels of a selection of six significant individual inflammatory genes: IL-6 (interleukin 6, A and G), IL-8 (interleukin 8, B and H), IL-10 (interleukin 10, C and I), IL-1RA (interleukin 1 receptor antagonist, D and J), GM-CSF (granulocyte-macrophage colony-stimulating factor, E and K), and TNF-α (tumor necrosis factor alpha, F and L). Cytokines/Chemokine expression values were used after Ln (*natural logarithmi*) transformation. Linear regression was performed for the two individual groups, survival (green) and death (red), and shown are the respective *r* (correlation coefficient) and *p* values of the regression lines. A *p* value < 0·05 was considered statistically significant, i.e. strong positive or negative correlation between testosterone and cytokine/chemokine expression.

## Discussion

Avian influenza A (H7N9) virus is of public health importance that has caused 1,567 laboratory-confirmed cases and 615 deaths since early 2013 in China[13]. Exposure to poultry was considered a predominant risk factor for H7N9 infections based on epidemiological and virological studies [14,15]. Although many aspects of H7N9 infection were studied, systematic analysis of the male bias of H7N9 avian influenza detected across five waves in China were lacking.

Several animal studies illustrated that sex hormones pose biological variables that may contribute to sex differences in influenza pathogenesis. But there are still large gaps regarding evidence from human cohorts. In this study, we retrospectively selected cohorts (18-49 years old and those ≥50 years of age) from laboratory-confirmed H7N9 (*n=*98) and seasonal influenza (*n=*53) cases, epidemiological linked poultry workers (*n=*108) and close contacts of H7N9 cases that were virus-negative (*n=*71). We systematically analyzed the association of the main estrogen (estradiol) and androgen (testosterone) levels and correlated to cytokine and chemokine levels known to be hallmarks of influenza disease outcome in both sexes.

We observed a strong suppression of testosterone levels in H7N9 infected men in both age groups (18-49 years and ≥50 years). One limitation of this study is that we have no information regarding potential comorbidities in these groups, such as diabetes type II or obesity, which are known to lower testosterone levels in men[16–19]. However, our data is in line with data reported in murine models, where infection of healthy male mice with influenza A virus was shown to reduce testosterone levels by independent laboratories[10] [12]. Interestingly, treatment of female and male mice with testosterone protected animals from lethal outcome[10]. However, testosterone treatment of elderly male mice was less effective [20]. These data strongly hint towards an active reduction of testosterone levels by influenza A virus infection by a yet unknown mechanism.

For chronic infections, such as HCV or HIV, it was repeatedly shown that these infections cause low testosterone levels likely due to constant inflammation in infected individuals[21,22]. In line with chronic conditions, such as type II diabetes, it is believed that chronic inflammatory diseases can cause low testosterone levels. Since testosterone plays key roles in not only fertility but also innate and adaptive immunity, one would hypothesize that all these “testosterone level” decreasing hits may increase vulnerability towards infections in general.

Elevated pro-inflammatory cytokines and chemokines, such as TNF-α, IL-8, IFN-α2, IFN-γ, IL-6, MCP-1, IP10, and anti-inflammatory cytokine IL-10 were detected in both male and female H7N9 cases without major statistically significant differences. However, testosterone levels were negatively associated with specific cytokines and chemokines, such as IL-6, and survival rates in H7N9 infected men. This is particularly interesting, since high IL-6 levels were repeatedly reported to be a poor prognostic marker for severe infections, including coronavirus disease 2019 [23]. Albeit the underlying mechanism causing testosterone deficiency is not known, there is increasing evidence that testosterone levels may modulate inflammatory cytokines[24–27]. It would be interesting to see whether other acute infections, such as the current pandemic Severe Acute Respiratory Syndrome Coronavirus 2(SARS-CoV-2), may also cause low testosterone levels.

Clearly, future investigations are required to unravel the underlying mechanism of low testosterone levels in H7N9 infected men. Increasing knowledge on the regulation of sex hormones in acute virus infections will provide an important basis to understand diseases with sex disparity and to develop sex specific therapies against viral infections.

## Data Availability

All generated data is available in this manuscript. No external repository was used.

## Author contributions

GG, YLS and SSB conceived, designed and overviewed the experiments. YLS and DYW organized and coordinated the field investigations. TB and YKC collected and organized specimens. SB and TB established testosterone and estradiol measurements. TB, YKC and SB conducted testosterone and estradiol measurements. YKC conducted cytokine and chemokine measurements and real-time RT PCR on outpatients for seasonal influenza viruses. JD performed real-time RT PCR on close contacts of H7N9 patients and poultry workers. TC, JY, LJW and DYW provided and overviewed the epidemiology information from study participants. TB and SSB analyzed the data. SB, TB and SSB developed all figures. TB and GG wrote the manuscript. All authors revised the manuscript.

## Acknowledgement

We thank Chinese National Influenza Surveillance Network and all staff at local CDCs in China for their contribution on sample collection and diagnosis for H7N9 and seasonal influenza A viruses. We would also like to thank staff at Sino-Japan Friendship Hospital for the measurement of testosterone and estradiol. Finally, we thank the study participants and their families.Figure Legends

